# Early identification of suboptimal responders to metformin in type 2 diabetes using long-term real-world HbA1c trajectories

**DOI:** 10.64898/2026.07.17.26357984

**Authors:** Eunsol Yang, Andrew Riselli, Fei Xu, Sneha B. Sridhar, Mark Kvale, Kathleen M. Giacomini, Monique M. Hedderson, Sook Wah Yee, Rada M. Savic

**Affiliations:** Department of Bioengineering and Therapeutic Sciences, University of California, San Francisco, 1700 Fourth Street, San Francisco, CA 94158, United States of America; Division of Research, Kaiser Permanente Northern California, 4480 Hacienda Drive, Pleasanton, CA 94588, USA; Institute for Human Genetics, University of California, San Francisco, 513 Parnassus Avenue, San Francisco, CA 94143, United States of America

**Keywords:** Type 2 diabetes, Metformin, HbA1c trajectories, Treatment response, Phenotype stratification, Real-world data

## Abstract

**Aims:** Metformin remains the primary treatment for type 2 diabetes, yet over 40% of patients fail to maintain glycaemic control. We aimed to identify patients unlikely to respond to metformin prior to treatment initiation and to evaluate whether on-treatment management can improve glycaemic outcomes in suboptimal responders, informing early treatment decisions.

**Materials and Methods:** We analyzed 59,881 longitudinal HbA1c measurements from 7,105 patients with type 2 diabetes receiving metformin monotherapy using real-world electronic health records from Kaiser Permanente Northern California with up to six years of follow-up. We integrated demographic, clinical, genetic, and pharmacological factors to characterize metformin responder phenotypes and quantify the impact of adherence and weight control on time to glycaemic failure.

**Results:** Three distinct trajectory-based phenotypes were identified: good (63.6%), poor (8.9%), and non-responders (27.5%). Poor responders initially achieved glycaemic targets but lost control within 2.5 years, while non-responders showed minimal HbA1c reduction and failed within 1 year. Five baseline factors—HbA1c, age at diagnosis, body mass index, sex, and estimated glomerular filtration rate—classified phenotypes with good discrimination (area under the receiver operating characteristic curve = 0.84). Incorporating on-treatment HbA1c further enhanced identification of non-responders. Among suboptimal responders, weight control and improved adherence delayed glycaemic failure by approximately 7 months; however, eventual glycaemic failure remained likely.

**Conclusions:** We characterized three clinically relevant metformin responder phenotypes and showed that suboptimal responders can be identified early using baseline features. Poor and non-responders are unlikely to achieve durable glycaemic control with metformin alone and may require alternative treatment strategies.

## 1. Introduction

Diabetes is among the fastest-increasing chronic diseases worldwide and represents a major public health challenge, affecting an estimated 539 million adults as of 2024, with over 90% diagnosed with type 2 diabetes.^1^ Type 2 diabetes is a complex and heterogeneous condition with substantially varying drug responses and disease progression, highlighting the importance of understanding this variability for improved management.^2,3^

Metformin has been the cornerstone of the treatment for type 2 diabetes for decades and remains one of the most widely prescribed oral antidiabetic drugs globally, owing to its efficacy, safety, and cost.^4,5^ Nevertheless, response to metformin is highly variable, with studies reporting that more than 40% of patients on metformin monotherapy experience treatment failure, posing a significant concern given the well-established link between inadequate glycaemic control and an increased risk of diabetes-related complications and mortality.^6–8^ Therefore, early identification of individuals at risk of suboptimal metformin response is essential to inform better strategies and ultimately enhance clinical outcomes. Delayed recognition of suboptimal response may lead to prolonged exposure to ineffective therapy, thereby increasing the risk of preventable complications.

There have been ongoing efforts to characterize suboptimal responders to metformin by linking treatment outcomes with demographic, clinical, or genomic markers. However, most studies have focused on early glycaemic response, relying on single post-treatment HbA1c values or short-term changes^8–11^ rather than longitudinal response patterns,^12^ limiting the ability to capture the heterogeneous and dynamic nature of type 2 diabetes. Although recent research analyzing HbA1c trajectories has advanced understanding of diabetes heterogeneity and treatment response, these approaches have typically included patients on diverse antidiabetic therapies, which restricts insights into drug-specific effects.^13–16^ Comprehensive identification of patient phenotypes specifically linked to long-term metformin response and associated determinants remains limited. Furthermore, although improved medication adherence and weight reduction have been consistently associated with better glycaemic outcomes in type 2 diabetes,^17–19^ the magnitude of long-term glycaemic benefits achievable through these on-treatment factors remains poorly quantified, particularly across phenotypically distinct subgroups.

In this study, we leveraged real-world longitudinal HbA1c trajectories from patients with type 2 diabetes receiving metformin monotherapy to characterize clinically relevant phenotypes of metformin response and their predictors that enable identification before treatment initiation, and to assess the impact of on-treatment management on glycaemic outcomes across phenotypes, thereby informing evidence-based early treatment decisions.

## 2. Material and Methods

### 2.1. Data and Patient Population

We evaluated longitudinal HbA1c trajectories for up to six years following metformin initiation in patients with type 2 diabetes receiving metformin monotherapy. Data were obtained from electronic health records of Kaiser Permanente Northern California (extracted in 2022), including laboratory measurements and pharmacy data. Patients were included if diagnosed at age ≥30 years to minimize inclusion of type 1 diabetes. HbA1c measurements obtained after initiation of any additional antidiabetic medications were excluded. Patients were eligible if they had received metformin monotherapy for at least three months, had a final HbA1c measurement recorded more than 90 days after metformin initiation, and had at least two HbA1c measurements. Included patients had no history of other antidiabetic medications for at least two years prior to metformin initiation.

### 2.2. Genotyping

Genotypes were derived from samples that had been genotyped and published in previous studies.^20–22^ We extracted two genotypes for evaluation: an intronic single nucleotide polymorphism (SNP) in the *SLC2A2* gene (rs8192675) and a SNP near the *ATM* gene (rs11212617), both of which have previously been reported to reach genome-wide significance for metformin response within 18 months of treatment.^22,23^

### 2.3. Longitudinal HbA1c Analysis

#### 2.3.1. Model development

We developed a longitudinal HbA1c model incorporating initial response and progression during metformin using a nonlinear mixed-effect modelling approach with NONMEM (version 7.5.1, ICON Development Solutions, Ellicott City, United States). To account for heterogeneity in glycaemic response and to stratify HbA1c trajectories, we applied a probabilistic mixture model based on parameters describing initial response and progression.^24^ The final model included two subpopulations for baseline HbA1c and initial response (low vs. high), and two for progression (slow vs. rapid), resulting in four distinct trajectories. Patients were assigned to the most likely trajectory based on the highest individual posterior probability estimated from observed HbA1c data.

#### 2.3.2. Phenotype classification

Each trajectory was defined as a metformin responder phenotype—good responders with faster or slower glycaemic control, poor responders, and non-responders—based on the extent of initial HbA1c reduction and time to loss of glycaemic control. The glycaemic goal was defined as HbA1c <7.0% (<53 mmol/mol) according to the 2026 American Diabetes Association guidelines.^25^

#### 2.3.3. Determinant identification

Significant determinants of initial response and progression in the longitudinal HbA1c model were evaluated stepwise, starting with baseline demographic and clinical factors, followed by genotypes, and then on-treatment variables. Variable selection was guided by statistical significance, clinical relevance, and prior evidence. Forward selection and backward elimination were conducted using significance thresholds of p<0.05 and p<0.01, respectively. The model building process was guided by numerical and graphical assessments, including the likelihood ratio test and visual predictive checks. Details of longitudinal HbA1c model analysis are provided in the Supplemental Methods.

### 2.4. Patient Classification Model Development

We developed multinomial logistic regression models using the nnet package in R (version 4.2.3) to classify patients into responder phenotypes assigned by the longitudinal HbA1c model. Candidate predictors included variables associated with initial response and progression identified in the longitudinal analysis. Patients with missing predictor values were excluded. Two primary models were built sequentially: (1) baseline characteristics only and (2) baseline plus on-treatment characteristics. Predictor selection followed the same stepwise procedure as in the longitudinal HbA1c analysis.

### 2.5. Longitudinal Model-Based Simulation

We applied the longitudinal HbA1c model to simulate HbA1c dynamics for up to six years after metformin initiation. For baseline characteristics, we used the ‘Base + Baseline’ model and simulated 1,000 virtual patients per determinant using 5^th^ or 95^th^ percentile values, with other factors held at median or reference values from our dataset, alongside 1,000 typical patients with all determinants set to median or reference values. For genotypes and on-treatment management, we used the ‘Base + Baseline + Genotype + On-treatment’ model and simulated 7,105 real-world patients per group using individual-level baseline characteristics and assigned phenotypes from our dataset.

## 3. Results

### 3.1. Data Characteristics

A total of 59,881 HbA1c measurements from 7,105 patients with type 2 diabetes receiving metformin monotherapy were included in the longitudinal analysis, with a median observation period of 4.2 years (Table 1). The study population had a balanced sex distribution with 51% male and was predominantly White (70.5%), with 10.4% Black and 6.8% Asian. At metformin initiation, baseline HbA1c and diabetes duration were widely distributed, with medians (2.5th to 97.5th percentiles) of 7.5% (6.0 – 12.8%) [58 (42 – 116 mmol/mol)] and 0.9 years (0.0 – 10.2 years), respectively. The *SLC2A2* rs8192675 C allele frequency was 0.32, with <2% missingness. The median average metformin daily dose was 1,000 mg, and the average adherence varied substantially between patients, with a median of 96% (2.5th to 97.5th percentiles: 20 – 100%).

**Table 1.**
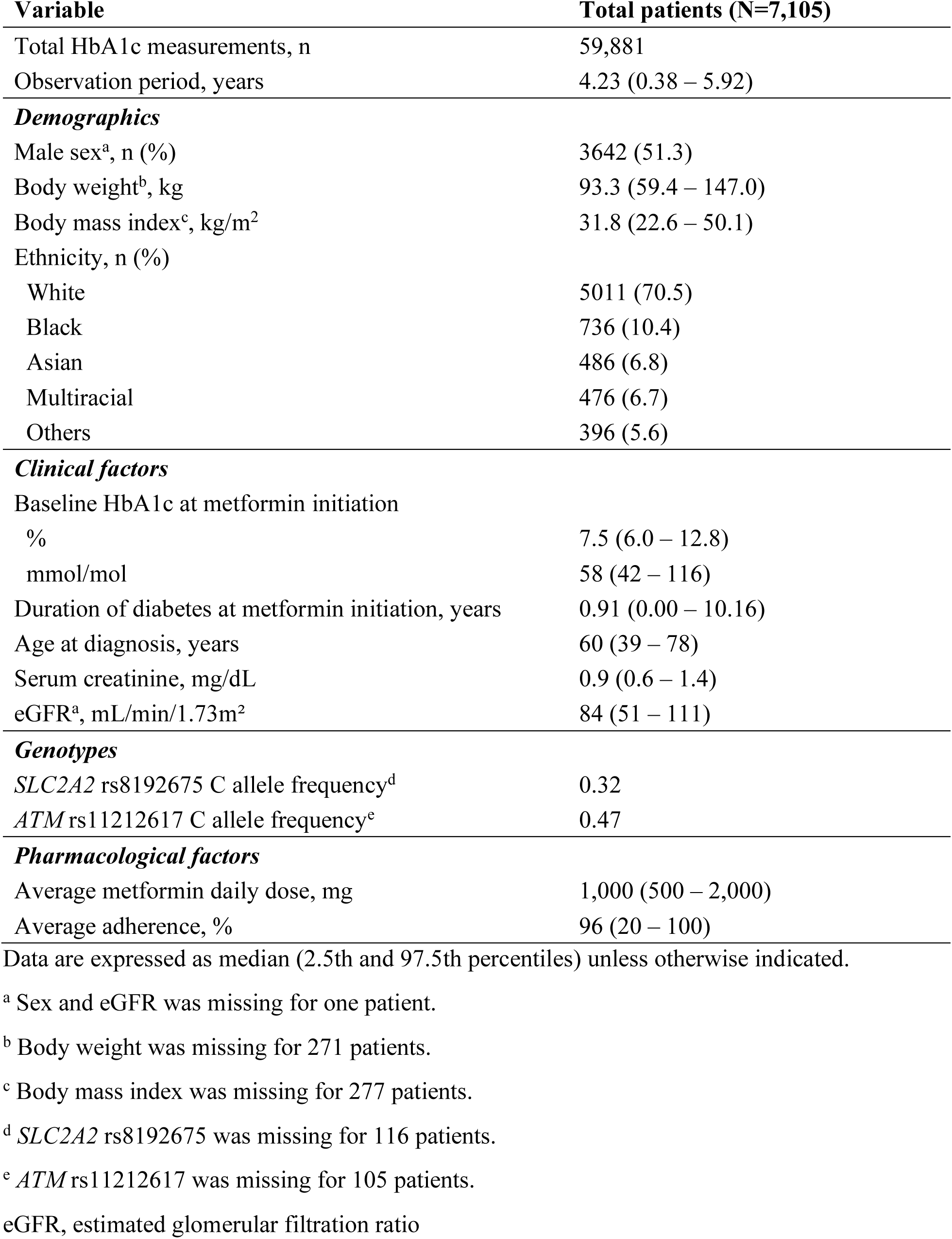
Demographic, clinical, genetic, and pharmacological characteristics of patients with type 2 diabetes.

| Variable | Total patients (N=7,105) |
| --- | --- |
| Total HbA1c measurements, n | 59,881 |
| Observation period, years | 4.23 (0.38 – 5.92) |
| ***Demographics*** |  |
| Male sex<sup>a</sup>, n (%) | 3642 (51.3) |
| Body weight<sup>b</sup>, kg | 93.3 (59.4 – 147.0) |
| Body mass index<sup>c</sup>, kg/m<sup>2</sup> | 31.8 (22.6 – 50.1) |
| Ethnicity, n (%) |  |
| White | 5011 (70.5) |
| Black | 736 (10.4) |
| Asian | 486 (6.8) |
| Multiracial | 476 (6.7) |
| Others | 396 (5.6) |
| ***Clinical factors*** |  |
| Baseline HbA1c at metformin initiation |  |
| % | 7.5 (6.0 – 12.8) |
| mmol/mol | 58 (42 – 116) |
| Duration of diabetes at metformin initiation, years | 0.91 (0.00 – 10.16) |
| Age at diagnosis, years | 60 (39 – 78) |
| Serum creatinine, mg/dL | 0.9 (0.6 – 1.4) |
| eGFR<sup>a</sup>, mL/min/1.73m<sup>2</sup> | 84 (51 – 111) |
| ***Genotypes*** |  |
| *SLC2A2* rs8192675 C allele frequency<sup>d</sup> | 0.32 |
| *ATM* rs11212617 C allele frequency<sup>e</sup> | 0.47 |
| ***Pharmacological factors*** |  |
| Average metformin daily dose, mg | 1,000 (500 – 2,000) |
| Average adherence, % | 96 (20 – 100) |

Data are expressed as median (2.5th and 97.5th percentiles) unless otherwise indicated.
<sup>a</sup> Sex and eGFR was missing for one patient.
<sup>b</sup> Body weight was missing for 271 patients.
<sup>c</sup> Body mass index was missing for 277 patients.
<sup>d</sup> *SLC2A2* rs8192675 was missing for 116 patients.
<sup>e</sup> *ATM* rs11212617 was missing for 105 patients.
eGFR, estimated glomerular filtration ratio

### 3.2. Metformin Responder Phenotypes based on Longitudinal HbA1c Trajectories

Three distinct metformin response phenotypes were identified based on longitudinal HbA1c trajectories: good responders (63.6%), poor responders (8.9%), and non-responders (27.5%) (Figure 1A–B). Longitudinal HbA1c data were best characterized using a turnover model incorporating separate effects of initial response and progression on the synthesis rate of HbA1c. The data were initially stratified into four distinct trajectories, including two subgroups of good responders with faster or slower glycaemic control (Supplemental Figure 1), which were subsequently combined given only marginal differences in time to glycaemic control (1 month for faster vs. 2.5 months for slower).

**Figure 1.**
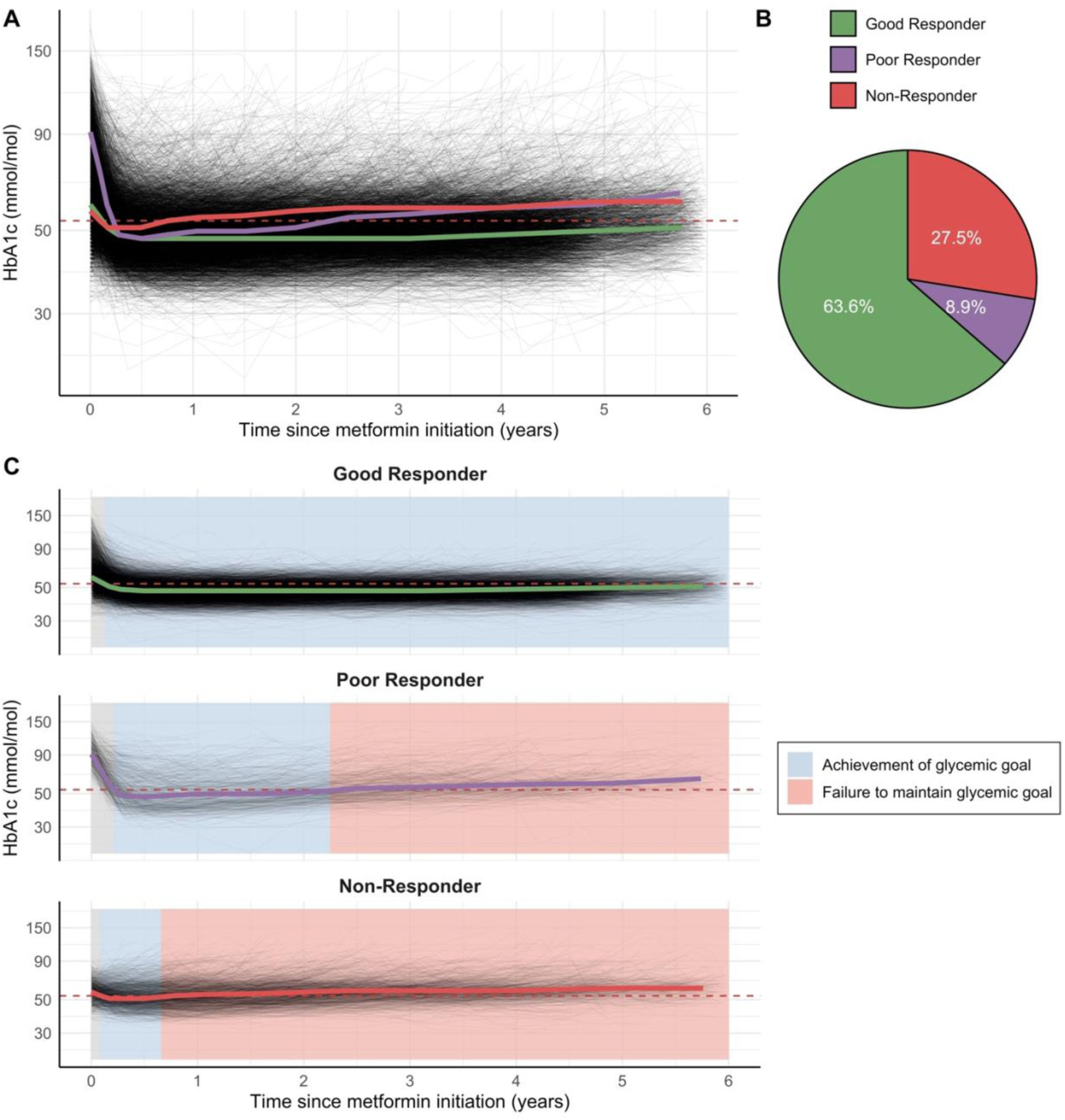
Longitudinal HbA1c trajectories and responder phenotype distribution under metformin monotherapy in patients with type 2 diabetes. (A) Individual HbA1c trajectories over six years with median lines by metformin responder phenotype. (B) Proportion of patients classified into each metformin responder phenotype. (C) Individual HbA1c trajectories over six years, highlighted by achievement of glycaemic goals (blue) and failure to maintain glycaemic control (red), stratified by metformin responder phenotype. The y-axis is log-scaled. The red dashed line represents the recommended HbA1c goal of <7.0% (<53 mmol/mol) according to the 2026 American Diabetes Association guidelines.

Poor responders initially achieved the recommended HbA1c goal of <7.0% (<53 mmol/mol) around six months after metformin initiation but failed to maintain this target within 2.5 years (Figure 1C). Non-responders showed minimal HbA1c reduction, with a median lowest HbA1c value of 6.8% (51 mmol/mol), and lost glycaemic control within approximately eight months of treatment. In contrast, good responders reached the HbA1c target within 2.5 months and sustained control for over six years under metformin monotherapy.

### 3.3. Patient Classification into Metformin Responder Phenotypes

A logistic regression model including five baseline features—HbA1c, age at diagnosis, body mass index (BMI), sex, and estimated glomerular filtration rate (eGFR)—classified patients into metformin responder phenotypes (Figure 2, Supplemental Table 1). Younger age at diagnosis, greater BMI, higher eGFR, and male sex were associated with a higher probability of poor or non-response. For example, the probability increased by 17% at 39 vs. 78 years of age, 13% at BMI 50.1 vs. 22.6 kg/m^2^, 7% in males vs. females, and 6% at eGFR 113 vs. 51 mL/min/m^2^.

**Figure 2.**
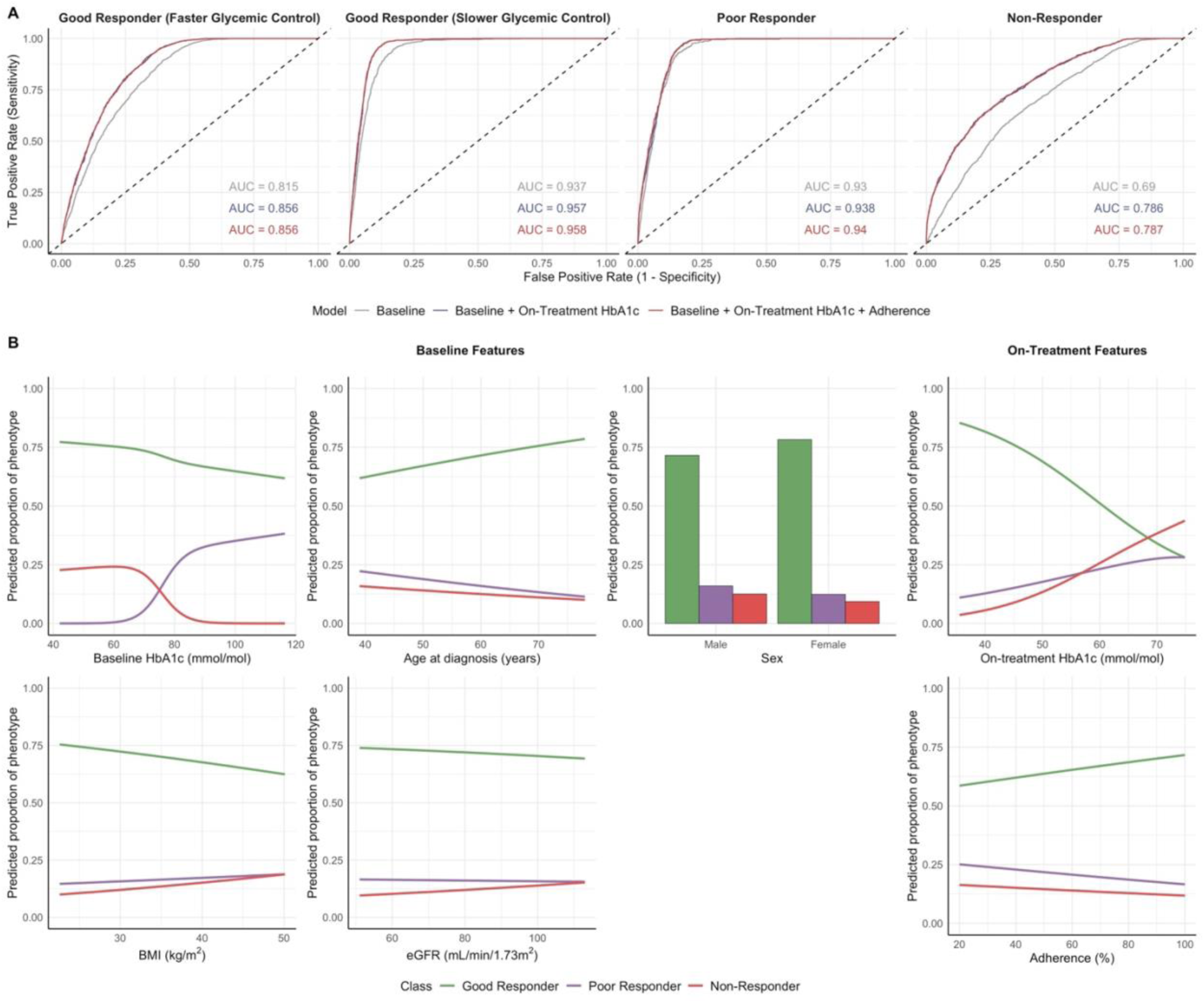
Logistic regression model for classifying patients into metformin responder phenotypes. (A) Receiver operating characteristic (ROC) curves evaluating the performance of logistic regression models. (B) Model-predicted proportions of metformin responder phenotypes based on individual baseline and on-treatment predictors. AUC, area under curve; BMI, body mass index; eGFR, estimated glomerular filtration ratio

Incorporating on-treatment HbA1c at around nine months after metformin initiation further improved overall phenotype discrimination, particularly for non-responder identification, with the area under the receiver operating characteristic curve (AUROC) increasing from 0.69 (95% confidence interval (CI), 0.68 – 0.71) to 0.79 (95% CI, 0.77 – 0.80) (Figure 2, Supplemental Table 2). Higher on-treatment HbA1c was strongly associated with poor or non-responder status, with 17% and 40% higher probabilities, respectively, in patients with HbA1c of 9.0% (75 mmol/mol) compared to 5.4% (36 mmol/mol).

Average adherence was also a significant predictor for phenotype classification; however, its inclusion only marginally improved model performance. Nonetheless, improved adherence mitigated the risk of poor or non-response, shifting patients toward the good responder phenotype. Specifically, increasing adherence from 20% to 100% was associated with a 13% absolute reduction in the probability of being classified as poor or non-responders.

### 3.4. Impact of Baseline Characteristics on Long-Term Glycaemic Outcomes

Greater BMI, higher eGFR, female sex, and Asian or Black race were significant determinants of lower initial response to metformin, while younger age at diagnosis, greater BMI, higher eGFR, and male sex were significant risk factors for more rapid progression during metformin monotherapy (Supplemental Table 3; ‘Base + Baseline’ model). When linked to HbA1c levels, these baseline predictors showed a less pronounced impact on glycaemic control in non-responders compared to other response phenotypes (Supplemental Figure 2). At six months, the predicted differences in median HbA1c levels for patients with each baseline risk factor relative to typical patients ranged from 0.0 to 0.3% in good responders, 0.2 to 0.5% in poor responders, and -0.1 to 0.1% in non-responders; corresponding ranges at six years were 0.1 to 0.3%, 0.2 to 0.5%, and -0.2 to 0.1%, respectively. Once phenotype was accounted for, variation in baseline risk factors did not influence whether each phenotype group was predicted to achieve the HbA1c goal of <7.0% (<53 mmol/mol).

### 3.5. Impact of Genotypes on Long-Term Glycaemic Outcomes

The *SLC2A2* rs8192675 C allele was significantly associated with greater initial metformin response, with response estimates 4.0% and 8.1% higher in the TC and CC groups, respectively, compared to the TT group (p < 0.001) (Supplemental Table 3; ‘Base + Baseline + Genotype’ model). However, it was not associated with progression during metformin treatment. When linked to HbA1c levels, *SLC2A2* genotypes showed a more notable impact on glycaemic control in poor responders and in good responders with slower glycaemic control compared to other phenotypes (Figure 3A). Compared to the TT group, the predicted improvement in HbA1c in the CC group at six months was 0.33% and 0.25% greater in poor responders and good responders with slower glycaemic control, respectively. In contrast, the corresponding differences were minimal in non-responders and in good responders with faster glycemic control (0.03% and 0.06%, respectively). The *ATM* rs11212617 C allele showed no significant association with either initial metformin response or progression.

**Figure 3.**
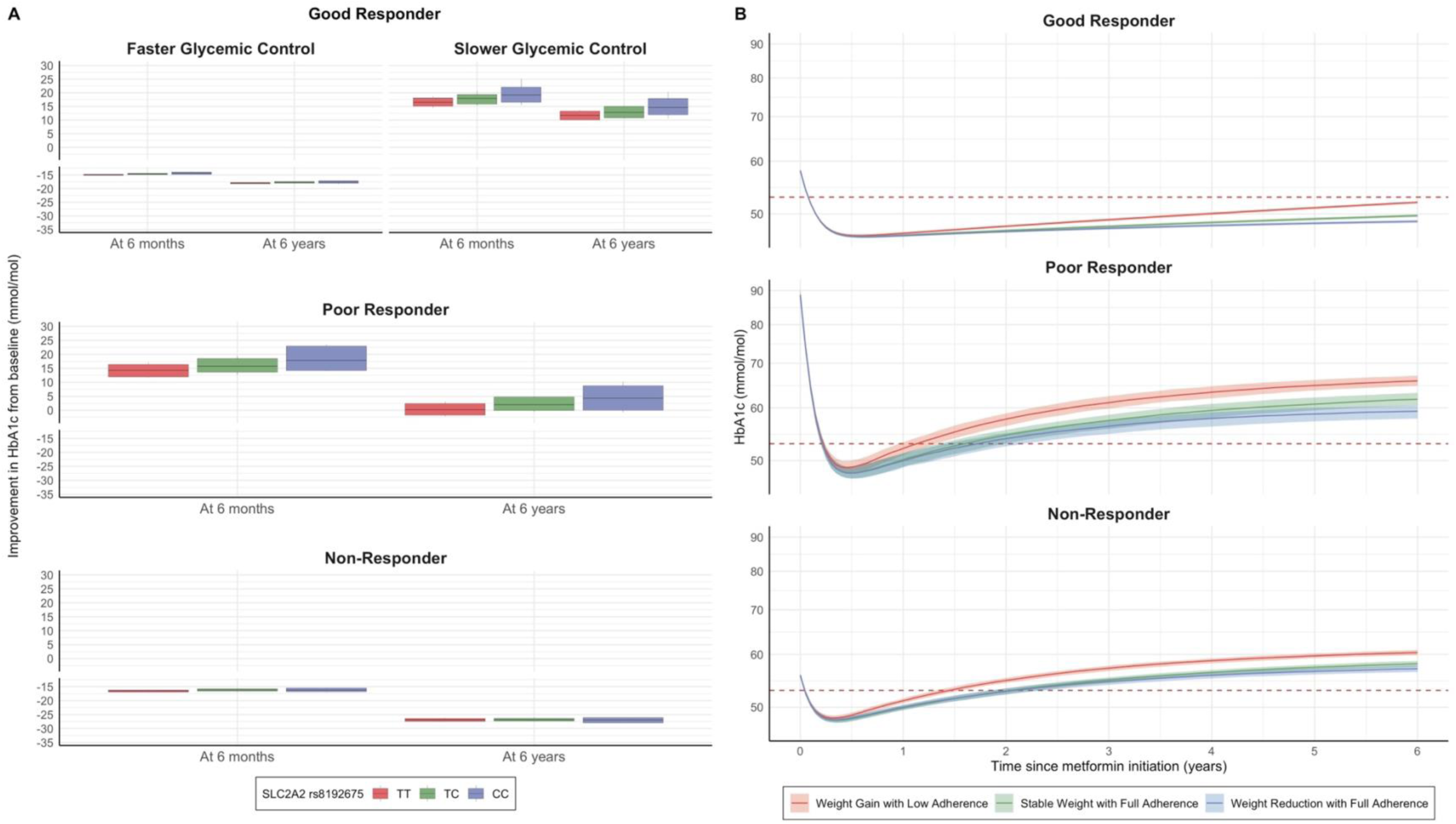
Effect of *SLC2A2* genotype and on-treatment management on long-term glycaemic outcomes by metformin responder phenotype. (A) Effect of the *SLC2A2* intronic variant (rs8192675 C allele) on HbA1c improvement from baseline at 6 months and 6 years after metformin initiation, stratified by metformin responder phenotype. The horizontal lines represent the medians of the simulated data. The boxes and vertical lines represent the 10^th^ and 90^th^ percentiles and minimum to maximum, respectively, of the simulated medians. (B) Effect of weight management and adherence on HbA1c trajectories, stratified by metformin responder phenotype. The solid lines represent the medians of the simulated data, and the shaded areas represent the 95% confidence intervals for the simulated medians. The red dashed line represents the recommended HbA1c goal of <7.0% (<53 mmol/mol) according to the 2026 American Diabetes Association guidelines.

### 3.6. Impact of On-Treatment Management on Long-Term Glycaemic Outcomes

Among on-treatment features, higher and increasing BMI, as well as lower and decreasing adherence, were significantly associated with more rapid progression on metformin (Supplemental Table 3; ‘Base + Baseline + Genotype + On-treatment’ model). In contrast, neither average metformin dose nor change in dose was associated with initial response or progression. When linked to HbA1c dynamics, weight management and improved adherence enhanced long-term glycaemic outcomes in suboptimal responders (Figure 3B). Compared to weight gain of 1.6% BMI per year with low adherence (33%, 5^th^ percentile), weight reduction of 1.6% BMI per year with full adherence (100%) delayed treatment failure by approximately seven months in poor and non-responders. These strategies also lowered predicted HbA1c levels at six years by up to 0.6% (7 mmol/mol) in poor responders and 0.3% (3 mmol/mol) in non-responders. However, regardless of on-treatment management, suboptimal responders were still expected to experience eventual glycaemic failure.

## 4. Discussion

We identified three distinct metformin responder phenotypes leveraging real-world longitudinal HbA1c trajectories. While the majority of patients (63.6%) were good responders who achieved and maintained the glycaemic goal over time, 36.4% exhibited suboptimal response characterized by more rapid progression towards treatment failure, comprising poor responders (8.9%) and non-responders (27.5%). Notably, non-responders had a more aggressive disease course than poor responders. Although non-responders tended to have lower baseline HbA1c values, their trajectories showed minimal initial improvement followed by rapid deterioration, indicating that these patterns were not solely driven by baseline glycaemic differences or floor effects. A recent analysis of two-year HbA1c trajectories in 140,413 patients with type 2 diabetes similarly identified three response patterns—stably low (89.7%), brisk response (6.6%), and non-response (3.1%)—but reported a substantially lower proportion of non-responders.^26^ This discrepancy may reflect the shorter follow-up period, which may not capture later glycaemic rebound or failure. In contrast, our work, despite a smaller cohort (n=7,105), included up to six years of follow-up, enabling more detailed classification of long-term glycaemic outcomes under metformin monotherapy.

Building upon the identified phenotypes, we developed classification tools to stratify individuals before or early during metformin treatment. A composite of five simple baseline features, associated with metformin pharmacokinetics, pharmacodynamics, or diabetes progression,^27–31^ classified patients into phenotypes, although accuracy for identifying non-responders was slightly lower compared to other subgroups. Incorporating on-treatment HbA1c at ∼nine months improved non-responder identification, resulting in AUROCs ranging from 0.79 to 0.96 across phenotypes. As non-responders typically lost glycaemic control by approximately eight months, this early marker helped distinguish them from good responders. Compared to prior models predicting metformin response,^8–11^ our approach has two key advantages: (1) reliance on commonly measured predictors, supporting feasibility for real-world implementation, and (2) response definitions based on HbA1c trajectories rather than single time points, better reflecting the dynamic nature of type 2 diabetes and long-term glycaemic control. By enabling early stratification, our tools may support more personalized, proactive treatment decisions, potentially improving early glycaemic control, which is strongly linked with reduced complications and mortality.^7,32^ To our knowledge, this represents one of the first prediction tools for classifying metformin responders based on longitudinal HbA1c trajectories.

In particular, we quantified the effects of changes in adherence and BMI over time as modifiable factors influencing glycaemic trajectories in the suboptimal group, given that unmodifiable baseline and genetic factors are fixed at metformin initiation and largely determine phenotype assignment. We found that, despite benefiting from improved adherence and weight reduction, suboptimal responders ultimately experience glycaemic failure. These findings suggest that, while such strategies can improve glycaemic control, consistent with previous reports,^18^ they alone may be insufficient to attain optimal outcomes in poor and non-responders. With the emergence of newer antidiabetic agents and more personalized, patient-centric pharmacologic approaches,^5^ suboptimal responders may benefit from alternative therapies or earlier treatment intensification with add-on agents that have demonstrated favorable efficacy in this population.^33–35^ Our findings provide a quantitative rationale for improving pharmacologic strategies in metformin suboptimal responders, alongside promoting adherence and weight management.

Higher baseline eGFR was associated with poorer longitudinal metformin response, which aligns with previous findings linking higher eGFR to less favorable glycaemic outcomes.^36^ Since metformin is predominantly eliminated via renal excretion,^29^ higher eGFR could result in lower drug exposure and thereby reduced response. Because younger age is related to greater renal function,^37^ and we observed a moderate negative correlation between baseline eGFR and age at diagnosis in our dataset (r = -0.5), part of this association may reflect the interplay with age and diabetes duration. Although diabetes progression is linked to declining kidney function over time,^38^ baseline eGFR remained a stronger determinant of lower initial response and more rapid progression than time-varying eGFR in our longitudinal model, suggesting that this association is not largely confounded by disease progression.

Using genotype data with <2% missingness, we uncovered the quantitative contribution of the SNP rs8192675 C allele in the *SLC2A2* gene, which encodes the glucose transporter isoform GLUT2,^39^ to glycaemic control on metformin. This intron variant has previously been associated with enhanced metformin response within 1 to 1.5 years of treatment by our group and others,^22,40^ in line with our findings. To our knowledge, this analysis is among the first to assess its association with long-term metformin response over six years and to link its impact on HbA1c dynamics across responder phenotypes. Poor responders exhibited the most pronounced impact, with noncarriers showing up to 0.33% less HbA1c reduction at six months compared to CC carriers. This difference corresponds to roughly half the HbA1c-lowering effect observed with the addition of a dipeptidyl peptidase 4 (DPP-4) inhibitor or sodium-glucose cotransporter 2 (SGLT2) inhibitor,^35,41^ and is clinically meaningful given the established relationship between HbA1c reduction and diabetes complications.^6^ Considering this variant in clinical practice could be especially relevant for poor responders; however, the feasibility of genotype-guided treatment decisions requires further evaluation.

Meanwhile, we did not observe a significant association between the SNP rs11212617 C allele in the *ATM* gene, involved in DNA repair and cell cycle control,^42^ and long-term metformin response in our cohort. While a previous genome-wide association study reported a significant association between this variant and early treatment success, defined as achieving HbA1c below 7% (53 mmol/mol) within the first 18 months of treatment,^23^ our six-year analysis in a larger cohort suggests its influence may be limited to short-term glycaemic outcomes.

We applied a nonlinear mixed-effect modelling approach combined with mixture models to characterize heterogeneity in metformin response and identify key determinants. Unlike traditional trajectory-based analyses,^13–16^ this approach captures both population-level patterns and individual-level variability, enabling refined phenotype classification while integrating mechanistic insights. This is particularly relevant given the heterogeneous nature of glycaemic trajectories in type 2 diabetes, where understanding individual-level dynamics is crucial for patient-centric approaches and improved clinical outcomes. As real-world and clinical trial data continue to expand, the methodology established herein could also be extended to other antidiabetic medications, providing a framework for analyzing long-term treatment response and uncovering influential factors, thereby advancing personalized and precision medicine in type 2 diabetes.

Collectively, we developed an integrated framework combining phenotype classification with prediction of long-term glycaemic outcomes utilizing baseline and on-treatment features. A key limitation is the absence of external validation, and further studies in independent cohorts are warranted to assess generalizability before clinical implementation. While the glycaemic target used in this study was based on the 2026 American Diabetes Association guidelines, alternative HbA1c targets used in different healthcare settings would not be expected to affect the underlying phenotype structure, as the phenotypes were derived from longitudinal HbA1c trajectory patterns; however, the interpretation of time to glycaemic failure may be influenced by the threshold used. If validated, this framework could support clinical decision-making through a sequential approach: (1) identifying metformin responder phenotypes using simple baseline measures prior to treatment initiation; (2) predicting glycaemic trajectories based on baseline characteristics; and (3) estimating the impact of on-treatment management on long-term outcomes. This framework may provide a practical roadmap for treatment and intervention strategies when initiating metformin in patients with type 2 diabetes and could be extended to incorporate diverse populations and broader social determinants of health, providing additional insights into factors contributing to heterogeneity in long-term metformin response.

In our analysis of real-world longitudinal HbA1c trajectories with demographic, clinical, genetic, and pharmacological factors, we identified three clinically relevant metformin responder phenotypes, with over one-third of patients exhibiting suboptimal glycaemic outcomes. Patients could be classified into these phenotypes using simple baseline measures, enabling early identification of those unlikely to respond well to metformin before treatment initiation.

Importantly, suboptimal responders are likely to experience eventual glycaemic failure despite on-treatment management, indicating the need for alternative therapeutic approaches in poor and non-responders. Once validated, our framework may inform phenotype-guided treatment strategies.

## Data Availability

The data that support the findings of this study are available from the authors on reasonable request.

## Author Contributions

F.X., S.B.S., K.M.G., M.M.H., and S.W.Y. contributed to data collection. E.Y., A.R., S.W.Y., and R.M.S. contributed to the analysis plan. E.Y. and A.R. prepared the data for analysis. K.M.G. and S.W.Y. performed and contributed to the genotyping. E.Y. performed the analysis with support from A.R., M.K., S.W.Y., and R.M.S. E.Y. drafted the manuscript with critical editorial input from S.W.Y. and R.M.S. All authors critically revised the manuscript for important intellectual content and approved the final manuscript for publication.

## Acknowledgements

At the time of this work, A.R. was supported by the National Institute of General Medical Sciences of the National Institutes of Health under award number 5T32GM007546.

## Prior Presentation

The work presented in this manuscript was presented in part at the American Society for Clinical Pharmacology and Therapeutics 2024 Annual Meeting, held in Colorado Springs, USA, from March 27 to 29, 2024.

## References

1. International Diabetes Federation. IDF Diabetes Atlas, 11th edn. Brussels, Belgium. International Diabetes Federation. Published online 2025.

2. Leslie RD, Ma RCW, Franks PW, Nadeau KJ, Pearson ER, Redondo MJ. Understanding diabetes heterogeneity: key steps towards precision medicine in diabetes. Lancet Diabetes Endocrinol. 2023;11(11):848–860. doi:10.1016/S2213-8587(23)00159-6

3. Ahlqvist E, Storm P, Käräjämäki A, et al. Novel subgroups of adult-onset diabetes and their association with outcomes: a data-driven cluster analysis of six variables. Lancet Diabetes Endocrinol. 2018;6(5):361–369. doi:10.1016/S2213-8587(18)30051-2

4. Drzewoski J, Hanefeld M. The Current and Potential Therapeutic Use of Metformin-The Good Old Drug. Pharmaceuticals (Basel). 2021;14(2):122. doi:10.3390/ph14020122

5. American Diabetes Association Professional Practice Committee for Diabetes*. 9. Pharmacologic Approaches to Glycemic Treatment: Standards of Care in Diabetes-2026. Diabetes Care. 2026;49(Supplement_1):S183–S215. doi:10.2337/dc26-S009

6. Stratton IM, Adler AI, Neil HA, et al. Association of glycaemia with macrovascular and microvascular complications of type 2 diabetes (UKPDS 35): prospective observational study. BMJ. 2000;321(7258):405–412. doi:10.1136/bmj.321.7258.405

7. Svensson E, Baggesen LM, Johnsen SP, et al. Early Glycemic Control and Magnitude of HbA1c Reduction Predict Cardiovascular Events and Mortality: Population-Based Cohort Study of 24,752 Metformin Initiators. Diabetes Care. 2017;40(6):800–807. doi:10.2337/dc16-2271

8. Bielinski SJ, Yanes Cardozo LL, Takahashi PY, et al. Predictors of Metformin Failure: Repurposing Electronic Health Record Data to Identify High-Risk Patients. J Clin Endocrinol Metab. 2023;108(7):1740–1746. doi:10.1210/clinem/dgac759

9. Rashid M, Shahzad M, Mahmood S, Khan K. Variability in the therapeutic response of Metformin treatment in patients with type 2 diabetes mellitus. Pak J Med Sci. 2019;35(1):71–76. doi:10.12669/pjms.35.1.100

10. Villikudathil AT, Mc Guigan DH, English A. Exploring metformin monotherapy response in Type-2 diabetes: Computational insights through clinical, genomic, and proteomic markers using machine learning algorithms. Comput Biol Med. 2024;171:108106. doi:10.1016/j.compbiomed.2024.108106

11. Naja K, Anwardeen N, Al-Hariri M, Al Thani AA, Elrayess MA. Pharmacometabolomic Approach to Investigate the Response to Metformin in Patients with Type 2 Diabetes: A Cross-Sectional Study. Biomedicines. 2023;11(8):2164. doi:10.3390/biomedicines11082164

12. Goswami S, Yee SW, Xu F, et al. A Longitudinal HbA1c Model Elucidates Genes Linked to Disease Progression on Metformin. Clin Pharmacol Ther. 2016;100(5):537–547. doi:10.1002/cpt.428

13. Luo M, Lim WY, Tan CS, et al. Longitudinal trends in HbA1c and associations with comorbidity and all-cause mortality in Asian patients with type 2 diabetes: A cohort study. Diabetes Res Clin Pract. 2017;133:69–77. doi:10.1016/j.diabres.2017.08.013

14. Luo M, Tan KHX, Tan CS, Lim WY, Tai ES, Venkataraman K. Longitudinal trends in HbA1c patterns and association with outcomes: A systematic review. Diabetes Metab Res Rev. 2018;34(6):e3015. doi:10.1002/dmrr.3015

15. Laiteerapong N, Karter AJ, Moffet HH, et al. Ten-year hemoglobin A1c trajectories and outcomes in type 2 diabetes mellitus: The Diabetes & Aging Study. J Diabetes Complications. 2017;31(1):94–100. doi:10.1016/j.jdiacomp.2016.07.023

16. Lavikainen P, Chandra G, Siirtola P, et al. Data-Driven Identification of Long-Term Glycemia Clusters and Their Individualized Predictors in Finnish Patients with Type 2 Diabetes. Clin Epidemiol. 2023;15:13–29. doi:10.2147/CLEP.S380828

17. Lean ME, Leslie WS, Barnes AC, et al. Primary care-led weight management for remission of type 2 diabetes (DiRECT): an open-label, cluster-randomised trial. Lancet. 2018;391(10120):541–551. doi:10.1016/S0140-6736(17)33102-1

18. McAdam-Marx C, Bellows BK, Unni S, et al. Impact of adherence and weight loss on glycemic control in patients with type 2 diabetes: cohort analyses of integrated medical record, pharmacy claims, and patient-reported data. J Manag Care Spec Pharm. 2014;20(7):691–700. doi:10.18553/jmcp.2014.20.7.691

19. Nichols GA, Rosales AG, Kimes TM, Tunceli K, Kurtyka K, Mavros P. The Change in HbA1c Associated with Initial Adherence and Subsequent Change in Adherence among Diabetes Patients Newly Initiating Metformin Therapy. J Diabetes Res. 2016;2016:9687815. doi:10.1155/2016/9687815

20. Wu B, Yee SW, Xiao S, et al. Genome-Wide Association Study Identifies Pharmacogenomic Variants Associated With Metformin Glycemic Response in African American Patients With Type 2 Diabetes. Diabetes Care. 2024;47(2):208–215. doi:10.2337/dc22-2494

21. Dawed AY, Yee SW, Zhou K, et al. Genome-Wide Meta-analysis Identifies Genetic Variants Associated With Glycemic Response to Sulfonylureas. Diabetes Care. 2021;44(12):2673–2682. doi:10.2337/dc21-1152

22. Zhou K, Yee SW, Seiser EL, et al. Variation in the glucose transporter gene SLC2A2 is associated with glycemic response to metformin. Nat Genet. 2016;48(9):1055–1059. doi:10.1038/ng.3632

23. GoDARTS and UKPDS Diabetes Pharmacogenetics Study Group, Wellcome Trust Case Control Consortium 2, Zhou K, et al. Common variants near ATM are associated with glycemic response to metformin in type 2 diabetes. Nat Genet. 2011;43(2):117–120. doi:10.1038/ng.735

24. Carlsson KC, Savić RM, Hooker AC, Karlsson MO. Modeling subpopulations with the $MIXTURE subroutine in NONMEM: finding the individual probability of belonging to a subpopulation for the use in model analysis and improved decision making. AAPS J. 2009;11(1):148–154. doi:10.1208/s12248-009-9093-4

25. American Diabetes Association Professional Practice Committee for Diabetes*. 6. Glycemic Goals, Hypoglycemia, and Hyperglycemic Crises: Standards of Care in Diabetes-2026. Diabetes Care. 2026;49(Supplement_1):S132–S149. doi:10.2337/dc26-S006

26. Raghavan S, Liu W, Warsavage T, Phillips LS, Reusch JE, Caplan L. 1201-P: Glycemic Response Phenotypes on Metformin Monotherapy in Real-World Diabetes Care. Diabetes. 2022;71(Supplement_1):1201-P. doi:10.2337/db22-1201-P

27. Deng L, Jia L, Wu XL, Cheng M. Association Between Body Mass Index and Glycemic Control in Type 2 Diabetes Mellitus: A Cross-Sectional Study. Diabetes Metab Syndr Obes. 2025;18:555–563. doi:10.2147/DMSO.S508365

28. Sherifali D, Nerenberg K, Pullenayegum E, Cheng JE, Gerstein HC. The effect of oral antidiabetic agents on A1C levels: a systematic review and meta-analysis. Diabetes Care. 2010;33(8):1859–1864. doi:10.2337/dc09-1727

29. Graham GG, Punt J, Arora M, et al. Clinical pharmacokinetics of metformin. Clin Pharmacokinet. 2011;50(2):81–98. doi:10.2165/11534750-000000000-00000

30. Zhou K, Donnelly LA, Morris AD, et al. Clinical and genetic determinants of progression of type 2 diabetes: a DIRECT study. Diabetes Care. 2014;37(3):718–724. doi:10.2337/dc13-1995

31. Gibson AA, Cox E, Schneuer FJ, et al. Sex differences in risk of incident microvascular and macrovascular complications: a population-based data-linkage study among 25 713 people with diabetes. J Epidemiol Community Health. 2024;78(8):479–486. doi:10.1136/jech-2023-221759

32. Laiteerapong N, Ham SA, Gao Y, et al. The Legacy Effect in Type 2 Diabetes: Impact of Early Glycemic Control on Future Complications (The Diabetes & Aging Study). Diabetes Care. 2019;42(3):416–426. doi:10.2337/dc17-1144

33. Xie Z, Hu J, Gu H, Li M, Chen J. Comparison of the efficacy and safety of 10 glucagon-like peptide-1 receptor agonists as add-on to metformin in patients with type 2 diabetes: a systematic review. Front Endocrinol (Lausanne). 2023;14:1244432. doi:10.3389/fendo.2023.1244432

34. Alnuaimi S, Reljic T, Abdulla FS, et al. PPAR agonists as add-on treatment with metformin in management of type 2 diabetes: a systematic review and meta-analysis. Sci Rep. 2024;14(1):8809. doi:10.1038/s41598-024-59390-z

35. Xu L, Wu Y, Li J, et al. Efficacy and safety of 11 sodium-glucose cotransporter-2 inhibitors at different dosages in type 2 diabetes mellitus patients inadequately controlled with metformin: a Bayesian network meta-analysis. BMJ Open. 2025;15(2):e088687. doi:10.1136/bmjopen-2024-088687

36. Garvey WT, Cohen RM, Butera NM, et al. Association of Baseline Factors With Glycemic Outcomes in GRADE: A Comparative Effectiveness Randomized Clinical Trial. Diabetes Care. 2024;47(4):562–570. doi:10.2337/dc23-1782

37. Glassock RJ, Winearls C. Ageing and the glomerular filtration rate: truths and consequences. Trans Am Clin Climatol Assoc. 2009;120:419–428.

38. Qiao X, Cai Q, Luo Y, Guo L, Pan Q. Longitudinal Trajectories of Albuminuria and eGFR in Type 2 Diabetes Mellitus: Natural Progression of Diabetic Kidney Disease. J Diabetes Res. 2025;2025:9269085. doi:10.1155/jdr/9269085

39. Thorens B. GLUT2, glucose sensing and glucose homeostasis. Diabetologia. 2015;58(2):221–232. doi:10.1007/s00125-014-3451-1

40. Rathmann W, Strassburger K, Bongaerts B, et al. A variant of the glucose transporter gene SLC2A2 modifies the glycaemic response to metformin therapy in recently diagnosed type 2 diabetes. Diabetologia. 2019;62(2):286–291. doi:10.1007/s00125-018-4759-z

41. Gilbert MP, Pratley RE. GLP-1 Analogs and DPP-4 Inhibitors in Type 2 Diabetes Therapy: Review of Head-to-Head Clinical Trials. Front Endocrinol (Lausanne). 2020;11:178. doi:10.3389/fendo.2020.00178

42. Ueno S, Sudo T, Hirasawa A. ATM: Functions of ATM Kinase and Its Relevance to Hereditary Tumors. Int J Mol Sci. 2022;23(1):523. doi:10.3390/ijms23010523

