## Supplemental Material for "Early identification of suboptimal responders to metformin in type 2 diabetes using long-term real-world HbA1c trajectories"

### Supplemental Methods

#### Demographic, Clinical, Genetic, and Pharmacological Factors

We considered a range of factors potentially associated with longitudinal HbA1c trajectories during metformin monotherapy. Demographics included sex, baseline and time-varying body mass index (BMI), and ethnicity. Clinical factors included baseline HbA1c, duration of diabetes, age at diagnosis, baseline and time-varying estimated glomerular filtration ratio (eGFR), on-treatment HbA1c at ~ nine months after metformin initiation. Genetic factors included the *SLC2A2* rs8192675 C allele and *ATM* rs11212617 C allele. Pharmacological factors included average and time-varying metformin daily dose, average and time-varying adherence, imputed metformin exposure. Adherence was calculated as the total number of days metformin was supplied divided by number of days in the observation period. Imputed metformin exposure was calculated by dividing the average daily dose by eGFR.

#### Longitudinal HbA1c Model Development and Evaluation

All parameters were estimated using first-order conditional method with interaction (FOCEI) option. Data processing and visualization were carried out using R (version 4.2.3), and the model was evaluated using Perl-speaks-NONMEM (PsN; version 5.3.0, Uppsala University, Sweden).

We used a mechanistic turnover model to characterize longitudinal HbA1 trajectories, incorporating both initial response and progression on metformin components. Various structural models for progression were explored, including linear, power, and negative exponential functions of time. Duration of diabetes was incorporated into the progression component to account for glycemic deterioration prior to metformin initiation. Inter-individual variability was evaluated using the exponential error model for baseline HbA1c and initial response parameters and the linear error model for progression parameter. Residual unexplained variability was described using a proportional model.

The known heterogeneity of HbA1c trajectories under metformin treatment was investigated through the use of a probabilistic mixture model based on initial response and progression parameters. Due to bimodal distributions of these parameters and a strong correlation between baseline HbA1c and initial response to metformin in our dataset, the model was parameterized with two subpopulations for baseline HbA1c and initial response (low vs. high) and two subpopulations for progression (slow vs. rapid), resulting in a total of four distinct trajectories.

Each model selection was guided by numerical assessments, including the likelihood ratio test and the precision of parameter estimates. The predictive performance of the model was evaluated graphically using the visual predictive check, where the observations were compared with the 200 simulated HbA1c-time profiles.

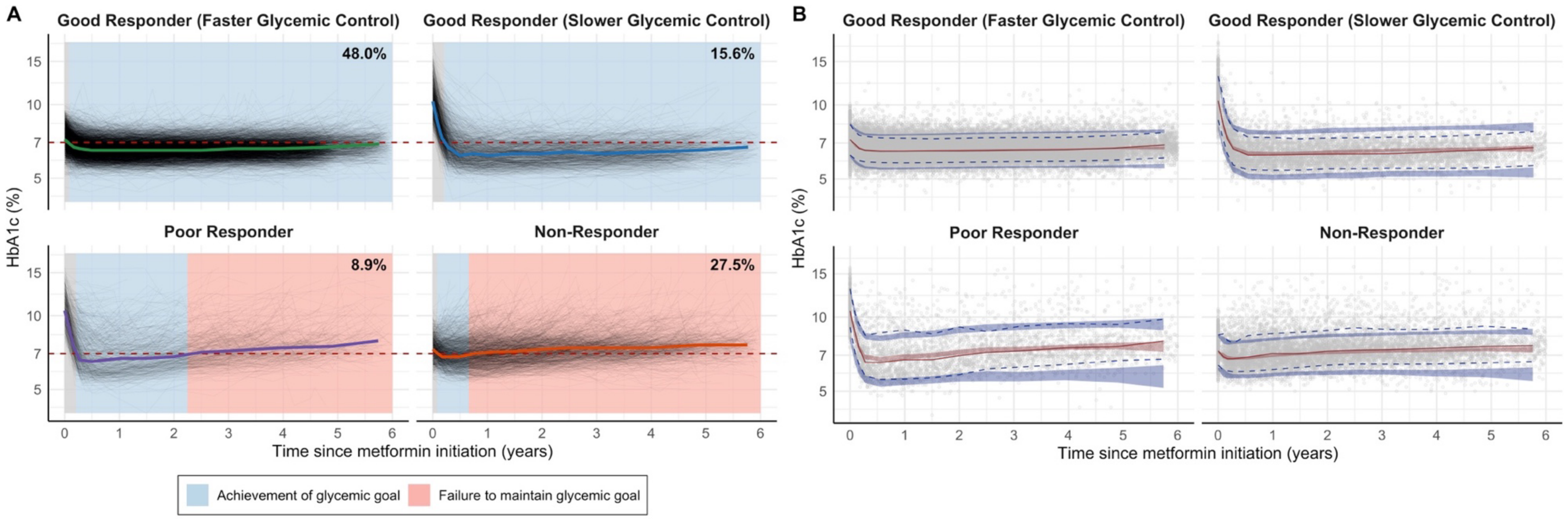


**Supplemental Figure 1.** Longitudinal HbA1c trajectories and model-based visual predictive check under metformin monotherapy in patients with type 2 diabetes.

(A) Individual HbA1c trajectories over 6 years, highlighted by achievement of glycemic goals (blue) and failure to maintain glycemic control (red), stratified by metformin responder phenotype. The red dashed line represents the recommended HbA1c goal of <7.0% (<53 mmol/mol) according to the 2026 American Diabetes Association guidelines.

(B) Visual predictive check of the longitudinal HbA1c model, stratified by metformin responder phenotype. The gray circles represent the observed HbA1c levels. The dashed lines represent the medians of the observed data. The shaded areas represent the 95% confidence intervals for the medians (red) and for the 10$^{th}$ and 90$^{th}$ percentiles (blue) of the simulated data.

The y-axis is log-scaled in both panels.

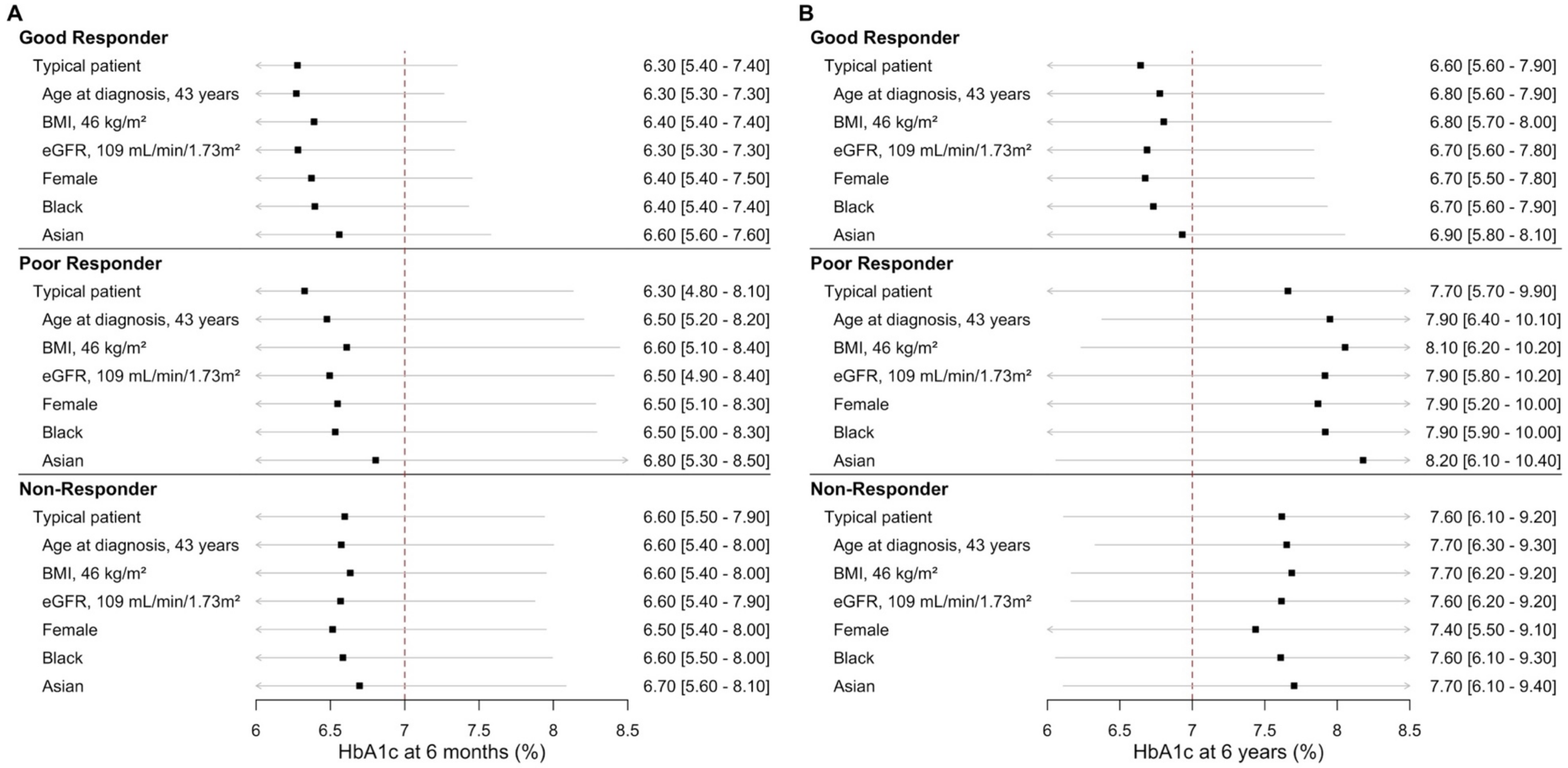


**Supplemental Figure 2.** Effect of baseline characteristics on HbA1c levels at (A) 6 months and (B) 6 years after metformin initiation, stratified by metformin responder phenotype.

A typical patient represents a hypothetical White male with median age at diagnosis, baseline BMI, and baseline eGFR.

The squares represent the medians of the simulated data. The gray lines represent the 95% confidence intervals for the simulated medians. The red dashed line represents the recommended HbA1c goal of <7.0% (<53 mmol/mol) according to the 2026 American Diabetes Association guidelines.

Abbreviations: BMI, body mass index; eGFR, estimated glomerular filtration rate

**Supplemental Table 1.** Baseline logistic regression model for classifying patients into metformin responder phenotypes.

| Variables | Univariable analysis | | | Multivariable analysis | | |
|---|---|---|---|---|---|---|
| | Good responder with slower glycemic control | Poor responder | Non-responder | Good responder with slower glycemic control | Poor responder | Non-responder |
| | vs. Good responder with faster glycemic control | | | vs. Good responder with faster glycemic control | | |
| | **OR [95% CI]** | **OR [95% CI]** | **OR [95% CI]** | **OR [95% CI]** | **OR [95% CI]** | **OR [95% CI]** |
| **Baseline Features** | | | | | | |
| ***Demographics*** | | | | | | |
| Female sex (relative to male) | 0.70 [0.61 - 0.80]*** | 0.51 [0.42 - 0.62]*** | 0.77 [0.68 - 0.87]*** | 0.74 [0.57 - 0.97]* | 0.53 [0.39 - 0.72]*** | 0.68 [0.59 - 0.78]*** |
| BMI (per 5-unit increase) | 1.07 [1.02 - 1.13]*** | 1.15 [1.07 - 1.22]*** | 1.21 [1.16 - 1.27]*** | 1.08 [0.98 - 1.18] | 1.15 [1.03 - 1.28]* | 1.17 [1.12 - 1.23]*** |
| Asian race (relative to White) | 0.60 [0.44 - 0.82]** | 0.55 [0.36 - 0.85]** | 0.58 [0.44 - 0.77]*** | - | - | - |
| Black race (relative to White) | 1.50 [1.22 - 1.84]*** | 1.46 [1.11 - 1.93]** | 1.04 [0.84 - 1.29] | - | - | - |
| Mixed race (relative to White) | 0.93 [0.71 - 1.23] | 0.51 [0.32 - 0.83]** | 0.85 [0.66 - 1.11] | - | - | - |
| ***Clinical factors*** | | | | | | |
| Baseline HbA1c (per 1-unit increase) | 15.8 [13.3 - 18.6]*** | 17.0 [14.3 - 20.3]*** | 1.19 [1.11 - 1.29]*** | 17.1 [14.2 - 20.5]*** | 18.6 [15.4 - 22.5]*** | 1.06 [0.98 - 1.16] |
| Age at diagnosis (per 10-unit increase) | 0.63 [0.58 - 0.67]*** | 0.49 [0.45 - 0.54]*** | 0.72 [0.68 - 0.77]*** | 0.82 [0.70 - 0.96]* | 0.64 [0.53 - 0.77]*** | 0.84 [0.78 - 0.91]*** |
| eGFR (per 10-unit increase) | 1.28 [1.23 - 1.34]*** | 1.40 [1.31 - 1.48]*** | 1.18 [1.13 - 1.22]*** | 1.00 [0.91 - 1.10] | 0.99 [0.89 - 1.10] | 1.10 [1.05 - 1.16]*** |

*** p-value < 0.001; **: p-value < 0.01* p-value < 0.05

Abbreviations: BMI, body mass index; CI, confidence interval; eGFR, estimated glomerular filtration ratio; OR, odds ratio

**Supplemental Table 2.** Baseline + On-treatment logistic regression model for classifying patients into metformin responder phenotypes.

| Variables | Univariable analysis | | | Multivariable analysis | | |
|---|---|---|---|---|---|---|
| | Good responder with slower glycemic control | Poor responder | Non-responder | Good responder with slower glycemic control | Poor responder | Non-responder |
| | vs. Good responder with faster glycemic control | | | vs. Good responder with faster glycemic control | | |
| | OR [95% CI] | OR [95% CI] | OR [95% CI] | OR [95% CI] | OR [95% CI] | OR [95% CI] |
| **Baseline Features** | | | | | | |
| ***Demographics*** | | | | | | |
| Female sex (relative to male) | 0.69 [0.60 - 0.79]*** | 0.56 [0.46 - 0.67]*** | 0.75 [0.66 - 0.85]*** | 0.93 [0.66 - 1.31] | 0.75 [0.52 - 1.09] | 0.7 [0.61 - 0.80]*** |
| BMI (per 5-unit increase) | 1.14 [1.09 - 1.20]*** | 1.04 [0.97 - 1.11] | 1.05 [1.01 - 1.10]* | 1.70 [1.50 - 1.92]*** | 1.40 [1.22 - 1.61]*** | 0.97 [0.93 - 1.02] |
| Asian race (relative to White) | 0.60 [0.44 - 0.82]** | 0.62 [0.41 - 0.93]* | 0.72 [0.56 - 0.92]** | - | - | - |
| Black race (relative to White) | 1.39 [1.13 - 1.72]** | 1.43 [1.09 - 1.87]** | 0.95 [0.77 - 1.18] | - | - | - |
| Mixed race (relative to White) | 0.92 [0.70 - 1.21] | 0.56 [0.36 - 0.87]** | 0.85 [0.66 - 1.09] | - | - | - |
| ***Clinical factors*** | | | | | | |
| Baseline HbA1c (per 1-unit increase) | 20.3 [16.9 - 24.5]*** | 22.2 [18.3 - 26.9]*** | 1.22 [1.13 - 1.31]*** | 2.31 [1.58 - 3.38]*** | 2.99 [2.06 - 4.33]*** | 1.68 [1.45 - 1.95]*** |
| Age at diagnosis (per 10-unit increase) | 0.61 [0.57 - 0.66]*** | 0.50 [0.46 - 0.55]*** | 0.73 [0.69 - 0.78]*** | 0.87 [0.71 - 1.07] | 0.66 [0.53 - 0.83]*** | 0.83 [0.77 - 0.90]*** |
| eGFR (per 10-unit increase) | 1.29 [1.24 - 1.35]*** | 1.39 [1.31 - 1.47]*** | 1.18 [1.14 - 1.22]*** | 0.93 [0.82 - 1.05] | 0.89 [0.78 - 1.02] | 1.10 [1.05 - 1.16]*** |
| **Genotypes** | | | | | | |
| *SLC2A2* rs8192675 TC (relative to TT) | 1.16 [1.00 - 1.35]* | 1.14 [0.93 - 1.38] | 0.95 [0.83 - 1.09] | - | - | - |
| *SLC2A2* rs8192675 CC (relative to TT) | 1.24 [1.00 - 1.54]* | 1.45 [1.10 - 1.89]** | 1.03 [0.85 - 1.25] | - | - | - |
| **On-treatment Features** | | | | | | |
| On-treatment HbA1c (per 1-unit increase) | 0.80 [0.72 - 0.88]*** | 1.88 [1.73 - 2.06]*** | 2.07 [1.93 - 2.22]*** | 0.0003 [0.0001 - 0.0005]*** | 0.0007 [0.0003 - 0.0014]*** | 8.86 [7.48 - 10.5]*** |

| | | | | | | |
|---|---|---|---|---|---|---|
| Average adherence (per 10-unit increase) | 1.07 [1.03 - 1.11]*** | 0.96 [0.93 - 1.00] | 0.92 [0.89 - 0.94]*** | 0.99 [0.91 - 1.07] | 0.91 [0.83 - 0.99]* | 0.94 [0.92 - 0.97]*** |
| ***Interaction*** | | | | | | |
| Baseline HbA1c: On-treatment HbA1c | - | - | - | 1.86 [1.74 - 1.98]*** | 1.79 [1.68 - 1.9]*** | 0.87 [0.85 - 0.89]*** |

*** p-value < 0.001; **: p-value < 0.01* p-value < 0.05

Abbreviations: BMI, body mass index; CI, confidence interval; eGFR, estimated glomerular filtration ratio; OR, odds ratio

**Supplemental Table 3.** Parameter estimates of the longitudinal HbA1c model.

| | Base | Base + Baseline | Base + Baseline + Genotype | Base + Baseline + Genotype + On-treatment |
|---|---|---|---|---|
| **Parameters** | **Estimates (RSE%)** | **Estimates (RSE%)** | **Estimates (RSE%)** | **Estimates (RSE%)** |
| Low baseline HbA1c, $HBBASE_{low}$ (%) | 7.28 (0.2%) | 7.28 (0.2%) | 7.24 (0.3%) | 7.22 (0.3%) |
| Low initial response, $MET_{low}$ | 0.119 (0.5%) | 0.119 (1.6%) | 0.118 (2.0%) | 0.118 (1.6%) |
| High baseline HbA1c, $HBBASE_{high}$ (%) | 10.3 (0.9%) | 10.3 (0.9%) | 10.2 (0.9%) | 10.2 (1.0%) |
| High initial response, $MET_{high}$ | 0.392 (1.3%) | 0.404 (1.4%) | 0.397 (1.6%) | 0.386 (1.6%) |
| Proportion of patients with $HBBASE_{low}$&$MET_{low}$ | 0.769 (1.3%) | 0.764 (1.4%) | 0.766 (1.4%) | 0.755 (1.5%) |
| Half-life of metformin response (day) | 31.4 (1.0%) | 31.3 (1.1%) | 31.4 (1.1%) | 31.3 (1.0%) |
| Slow progression, $DISPR_{slow}$ ($day^{-1}$) | 0.0149 (3.8%) | 0.0145 (4.2%) | 0.0147 (4.9%) | 0.0147 (3.7%) |
| Rapid progression, $DISPR_{rapid}$ ($day^{-1}$) | 0.135 (1.0%) | 0.109 (1.5%) | 0.111 (4.5%) | 0.0975 (1.6%) |
| Proportion of patients with $DISPR_{slow}$ | 0.723 (1.6%) | 0.668 (2.2%) | 0.671 (2.3%) | 0.636 (2.4%) |
| $DISPR_{max}$ | 0.202 FIX | 0.202 FIX | 0.202 FIX | 0.202 FIX |
| IIV on $HBBASE_{low}$ (CV%) | 9.6 (4.5%) | 9.6 (4.5%) | 9.5 (4.5%) | 9.3 (4.7%) |
| Correlation $HBBASE_{low}$ - $MET_{low}$ (%) | 32.5 (23.8%) | 32.3 (26.8%) | 33.3 (25.7%) | 34.9 (24.5%) |
| IIV on $MET_{low}$ (CV%) | 38.9 (14.0%) | 39.1 (14.6%) | 39.6 (14.1%) | 40.4 (13.1%) |
| IIV on $HBBASE_{high}$ (CV%) | 16.2 (2.9%) | 16.4 (2.9%) | 16.2 (3.1%) | 16.4 (3.9%) |
| Correlation $HBBASE_{high}$ - $MET_{high}$ (%) | 67.9 (7.6%) | 70.1 (7.0%) | 68.5 (7.4%) | 64.7 (8.0%) |
| IIV on $MET_{high}$ (CV%) | 22.5 (9.7%) | 22.5 (9.3%) | 22.2 (9.4%) | 24.5 (8.8%) |
| IIV on $DISPR_{slow}$ (CV%) | 94.9 (6.6%) | 69.0 (7.1%) | 68.0 (7.4%) | 68.0 (11.5%) |
| IIV on $DISPR_{rapid}$ (CV%) | 53.4 (13.5%) | 49.4 (9.4%) | 48.5 (9.9%) | 56.2 (7.6%) |
| Proportional residual error (%) | 7.78 (0.1%) | 7.78 (0.1%) | 7.78 (0.1%) | 7.67 (0.1%) |
| *Effect on baseline HbA1c* | | | | |
| Increase in *SLC2A2* rs8192675 TC (%) | – | – | 1.00 (35.3%) | 0.994 (35.3%) |
| Increase in *SLC2A2* rs8192675 CC (%) | – | – | 1.67 (31.5%) | 1.64 (31.8%) |

| | | | | |
|---|---|---|---|---|
| *Effect on initial response* | | | | |
| Baseline body mass index exponent | – | -0.113 (24.5%) | -0.120 (15.8%) | – |
| Time-varying body mass index exponent | – | – | – | -0.340 (5.7%) |
| Baseline glomerular filtration rate exponent | – | -0.0948 (29.5%) | -0.0957 (29.2%) | -0.0751 (39.1%) |
| Decrease in female (%) | – | 4.93 (20.1%) | 5.01 (20.2%) | 4.18 (25.4%) |
| Decrease in Asian relative to White (%) | – | 10.5 (19.2%) | 10.2 (19.9%) | 15.4 (12.8%) |
| Decrease in Black relative to White (%) | – | 4.22 (33.9%) | 5.77 (27.7%) | 5.09 (33.2%) |
| Increase in *SLC2A2* rs8192675 TC (%) | – | – | 3.31 (40.5%) | 3.98 (35.7%) |
| Increase in *SLC2A2* rs8192675 CC (%) | – | – | 7.47 (30.1%) | 8.06 (29.5%) |
| *Effect on progression* | | | | |
| Age at diagnosis exponent | – | -0.865 (12.8%) | -0.842 (10.1%) | -0.247 (42.5%) |
| Baseline body mass index exponent | – | 0.468 (22.6%) | 0.440 (4.0%) | – |
| Time-varying body mass index exponent | – | – | – | 2.07 (3.8%) |
| Baseline glomerular filtration rate exponent | – | 0.437 (29.7%) | 0.435 (28.0%) | 0.400 (28.7%) |
| Decrease in female (%) | – | 22.0 (14.9%) | 22.4 (14.8%) | 29.5 (9.1%) |
| Time-varying adherence exponent | – | – | – | -0.255 (5.1%) |

Abbreviations: CV, coefficient of variation; RSE, relative standard error
